# QUARTZ: Cross-Dataset Artificial Intelligence-Enabled Retinal Vasculometry

**DOI:** 10.64898/2026.10.28.26363792

**Authors:** Roshan A. Welikala, Jiri Fajtl, Gordon Johnson, Farzana Rahman, Paolo Remagnino, Ellen E. Freeman, Miriam Kolko, Ryan Chambers, Louis Bolter, John Anderson, Abraham Olvera-Barrios, Alasdair Warwick, Paul J. Foster, Anthony P. Khawaja, Royce Shakespeare, Misha Ramesh, Catherine Egan, Adnan Tufail, Christopher G. Owen, Alicja R. Rudnicka, Sarah Barman, the UK Biobank Eye and Vision Consortium, the EPIC-Norfolk Investigators, the ARIAS Research Group

**Author notes:** Corresponding author: Sarah Barman.

## Abstract

Examination of retinal images provides a non-invasive means to assess the body’s microvascular system, with vessel morphology linked to ocular and systemic diseases. QUARTZ (QUantitative Analysis of Retinal vessel Topology and siZe) is an automated, artificial intelligence-enabled retinal vasculometry system designed for processing large-scale retinal image datasets, including over 4 million images from within the National Health Service, to facilitate epidemiological studies. The system is uniquely capable of extracting useful information from lower quality images, including those which are partially illuminated, enabling analysis of a greater proportion of images. To enhance cross-dataset generalisation of the system from an earlier dataset-tailored approach, this work leverages 26 annotated retinal image datasets spanning diverse demographics, diseases, camera types, and imaging protocols. Evaluation on the external EPIC-Norfolk dataset demonstrates robust performance with F1 scores of 0.8305, 0.8477, 0.8741, 0.9532, and 0.8677 for arteriole, venule, vessel, optic disc, and optic cup segmentation, respectively, and 0.9197 for image quality classification. The cross-dataset generalisation approach demonstrated increased performance for arteriole/venule segmentation and optic disc segmentation. However, improvements in performance were not observed for optic cup segmentation, or for image quality classification. An online tool is provided to support wider adoption and reproducible biomarker research across diverse populations and imaging settings. In addition, the new public CHASE-AV database is introduced for arteriole and venule segmentation.

## (1) Introduction

Examination of retinal images provides a direct and non-invasive means to observe the body’s microvascular system. The morphological properties of retinal vessels, including their width, tortuosity, and branching pattern, have been prospectively linked to both ocular and systemic diseases [1,2]. As such, the retina serves as a valuable indicator of overall vascular health, offering potential biomarkers for early detection and risk prediction of conditions such as diabetic retinopathy, diabetes, hypertension, and cardiovascular disease [3].

The quantitative analysis of the retinal vasculature, or retinal vasculometry, has therefore become an important research tool. Despite advances in end-to-end deep learning for disease and risk prediction [4,5], accurate retinal vessel segmentation (the foundation of vasculometry) continues to be an active area of research [6,7]. Moreover, retinal vasculometry remains valuable because it yields interpretable, biologically grounded vascular features (e.g., width, tortuosity, branching geometry, and arteriole/venule differences) that can be directly linked to disease mechanisms and are amenable to epidemiological analysis and clinical interpretation. Several systems exist for this purpose, including AutoMorph [8], VascX [9], VAMPIRE [10], RHMAS [11], and QUARTZ [12] some of which have been developed as effective generalisation-focused tools capable of operating across multiple datasets.

Developing a generalised system [13–15] offers broad applicability across diverse populations and imaging settings but is challenged by heterogeneity across datasets. Variations in camera systems, field-of-view (FOV) angles, image resolution, image quality, pupil dilation (mydriasis), and ocular disease can influence deep learning models. Differences in population demographics, such as skin or hair colour, affect pigmentation and retinal appearance [16], potentially biasing models trained on homogenous datasets. Additionally, inconsistencies in annotation quality and styles across datasets can hinder model training and evaluation.

Importantly, a diverse combination of datasets needs to be utilised to enhance model generalisability [9] and robustness across heterogeneous imaging and population characteristics, encouraging the model to focus on intrinsic vascular features rather than dataset-specific variations. In addition, annotation standardisation is crucial to ensure consistency across datasets.

The approach of the vasculometry system QUARTZ (QUantitative Analysis of Retinal vessel Topology and siZe) has previously been reported. [12,17]. This automated artificial intelligence-enabled retinal vasculometry system. has been used to process several large retinal image datasets from cohort studies to obtain quantitative measures of vessel morphology. While other leading vasculometry systems demonstrate effective generalisation approaches [8,9,11], QUARTZ is uniquely optimised for epidemiological studies, with the ability to effectively perform segmentation and extract useful data from lower quality images (e.g., partially illuminated images) that are common in large cohort studies (reported in [12]). QUARTZ has enabled multiple epidemiological studies [3,18–27]. QUARTZ [12] consists of several deep learning models; however, it was separately tailored to specific datasets, namely the UK Biobank (UKBB) [28], the Canadian Longitudinal Study on Aging (CLSA) [29], and the North East London Diabetic Eye Screening Programme (NEL DESP) [30]. As imaging protocols become further standardised cross-dataset variability may be reduced, However substantial heterogeneity currently exists.

In this work, QUARTZ modules are trained on multiple datasets encompassing diverse demographics, a broad spectrum of diseases (from community to diseased populations), and varied camera setups and imaging protocols to enhance external generalisability. Attention is given to annotation standardisation across datasets to support stable and reliable model training. In addition, training includes images with varying and imperfect illumination to ensure reliable performance under such conditions. The system’s deep learning model performances are evaluated across all internal datasets to show model stability within known distributions and are additionally compared against the previous dataset-tailored approach. Results on multiple external datasets (unseen domains) are reported to demonstrate model generalisation. A move toward a generalisation-focused system can improve cross-dataset robustness, albeit with a potential reduction in performance compared with dataset-tailored systems, a pattern explored here.

The main contributions of this work are: (i) an assessment of the impact on performance of a generalisation- focused approach applied to the retinal vasculometry system QUARTZ, trained on multiple datasets with annotation standardisation performed and evaluated against dataset-tailored systems; (ii) an online tool is released (https://miqa.kingston.ac.uk/quartz/), supporting wider access and fostering reproducible biomarker research studies across diverse populations; (iii) a new public database, CHASE-AV, is introduced for arteriole/venule segmentation (https://researchinnovation.kingston.ac.uk/en/datasets/chase-av-retinal-vessel-reference-dataset/).

## (2) Materials

Numerous public and non-public annotated colour retinal image datasets were curated, totalling 26 datasets. Two datasets (EYEPACS [31] and EyeQ [32]) were used exclusively for pretraining; summary details for the remaining 24 annotated datasets are provided in Table 1. The non-public datasets, drawn from controlled-access cohort studies, had been previously annotated in another study [12]. The annotations, manually labelled by trained human observers, include pixel-level labels of arterioles and venules, pixel-level labels for the optic disc and cup, and image-level labels for image quality (adequate vs inadequate).

**Table 1:** Characteristics of the annotated retinal image datasets. Certain datasets, such as UKBB, are listed multiple times due to different annotated subsets corresponding to different label types. For UKBB, CLSA, NEL DESP, and EPIC-Norfolk, the original source of the retinal images is cited; the annotated datasets were derived later [12]. The symbol “∼” indicates an approximate value (not explicitly documented).

| Dataset | Label | No. of Images | Centred | FOV | Image Size (pixels) | Image format | Ocular Disease | Country | Camera |
| --- | --- | --- | --- | --- | --- | --- | --- | --- | --- |
| UKBB [28] (non-public) | arteriole/venule | 25 | macula | 45° | 2048 × 1536 | PNG | Low | UK | Topcon 3D-OCT 1000 Mark 2 |
| CLSA [29] (non-public) | arteriole/venule | 25 | macula | 45° | 4288 × 2848 | JPEG | age-related | Canada | Topcon TRC-NW8 |
| NEL DESP [30] (non-public) | arteriole/venule | 25 | macula, disc | 45° | 3648 × 2432 | JPEG | diabetic retinopathy | UK | Various Canon and Topcon models |
| RITE [35] (public) | arteriole/venule | 40 | macula, disc | 45° | 565 × 584 | TIFF | diabetic retinopathy | Netherlands | Canon CR5 3CCD |
| LES-AV [36] (public) | arteriole/venule | 22 | disc | 30°<br>45° | 1620 × 1444,<br>2196 × 1958 | PNG | glaucoma | - | Visucam Pro Nm |
| HRF-AV [33] (public) | arteriole/venule | 45 | macula | 60° | 3504 × 2336 | JPEG | diabetic retinopathy, glaucoma | Czech Republic | Canon CF-60 UVi |
| Leuven-Haifa [37] (public) | arteriole/venule | 240 | disc | 30° | 1444 × 1444 | PNG | glaucoma | Belgium | Zeiss Visucam 500 |
| CHASE-AV (public) | arteriole/venule | 28 | disc | 30° | 999 × 960 | JPEG | none | UK | hand-held Nidek NM-200-D |
| FOREVER [38,39] (non-public) | arteriole/venule | 20 | macula | 45° | 3984 × 2656 | BMP | low | Denmark | Canon CR-2 AF |
| Fundus-AVSeg [40] (public) | arteriole/venule | 100 | macula, disc | ~45° | 2656 × 1992,<br>1280 × 1280 | PNG | diabetic retinopathy, glaucoma, AMD | China | Zeiss Visucam200 + Canon models |
| EPIC-Norfolk [41] (non-public) | arteriole/venule | 25 | macula | 45° | 3008 × 2000 | JPEG | low | UK | Topcon TRC-NW65 |
| UKBB [28] (non-public) | optic disc/cup | 100 | macula | 45° | 2048 × 1536 | PNG | low | UK | Topcon 3D-OCT 1000 Mark 2 |
| CLSA [29] (non-public) | optic disc/cup | 100 | macula | 45° | 4288 × 2848 | JPEG | age-related | Canada | Topcon TRC-NW8 |
| NEL DESP [30] (non-public) | optic disc/cup | 100 | macula, disc | 45° | 3648 × 2432 | JPEG | diabetic retinopathy | UK | Various Canon and Topcon models |
| REFUGE [42] (public) | optic disc/cup | 1200 | macula | 45° | 2124 × 2056,<br>1634 × 1634 | JPEG | glaucoma | China | Zeiss Visucam 500 + Canon CR-2 |
| PAPILA [43] (public) | optic disc/cup | 488 | disc | 30° | 2576 × 1934 | JPEG | glaucoma | Spain | Topcon TRC-NW400 |
| Cháksu [44] (public) | optic disc/cup | 1345 | macula, disc | 40° | 2448 × 3264,<br>2048 × 1536,<br>1920 × 1440 | JPEG, PNG | glaucoma | India | Remidio Fundus-on-Phone + Forus 3Nethra Classic + Bosch handheld |
| ORIGA [45] (public) | optic disc/cup | 650 | macula, disc | ~45° | various | JPEG | glaucoma | Singapore | - |
| EPIC-Norfolk [41] (non-public) | optic disc/cup | 100 | macula | 45° | 3008 × 2000 | JPEG | low | UK | Topcon TRC-NW65 |
| UKBB [28] (non-public) | image quality | 2000 | macula | 45° | 2048 × 1536 | PNG | low | UK | Topcon 3D-OCT 1000 Mark 2 |
| CLSA [29] (non-public) | image quality | 2000 | macula | 45° | 4288 × 2848 | JPEG | age-related | Canada | Topcon TRC-NW8 |
| NEL DESP [30] (non-public) | image quality | 2000 | macula, disc | 45° | 3648 × 2432 | JPEG | diabetic retinopathy | UK | Various Canon and Topcon models |
| FQS [46] (public) | image quality | 2246 | macula, disc | ~45°<br>30° | 2304 × 1728 | PNG | various | China | Zeiss Visucam200 + Canon models |
| EPIC-Norfolk [41] (non-public) | image quality | 2000 | macula | 45° | 3008 × 2000 | JPEG | low | UK | Topcon TRC-NW65 |

The datasets cover a broad range of imaging conditions, including cameras ranging from smartphone-based devices to full retinal cameras, varying resolutions, FOV angles (30°–60°), and differences in image quality, such as illumination levels (including partially illuminated images). The retinal images represent diverse populations, with variation in age from children to older adults and across different ethnicities. Images were collected in a variety of clinical settings, ranging from high street opticians to hospital appointments. The datasets include subjects with a range of disease statuses, including ocular diseases such as diabetic retinopathy, age related macular degeneration, and glaucoma, as well as systemic conditions like cardiovascular disease.

Whilst pathological cases are included, vessel annotations do not extend to shunt vessels or neovascularisation. The NEL DESP [30] and HRF-AV [33] images are mydriatic, whereas the remaining datasets are non-mydriatic.

In addition, we introduce a new annotated public database, CHASE-AV, which provides pixel-level arteriole and venule labels. CHASE-AV is built upon the publicly available CHASE-DB1 [34] dataset and contains 28 retinal images along with corresponding annotation masks produced by an expert human observer with clinical experience. (https://researchinnovation.kingston.ac.uk/en/datasets/chase-av-retinal-vessel-reference-dataset/).

## (3) Methods

The QUARTZ pipeline was structured into five modules: pre-processing, arteriole/venule segmentation, optic disc/cup segmentation, image quality classification, and vessel morphometric analysis (see Figure 1). Three modules were based on deep learning models, and brief architectural details are provided in this section (see [12] for full details, including architectural design and justification). The deep learning modules mitigate image size variability through cropping and resizing. In addition, building a generalisation-focused system required annotation standardisation and joint training across multiple datasets. Training on multiple datasets was used to enhance external generalisability. The network architecture and data augmentation were kept consistent with the previous pipeline, while hyperparameters were tuned using the same search strategy to ensure a fair comparison.

**Figure 1:**
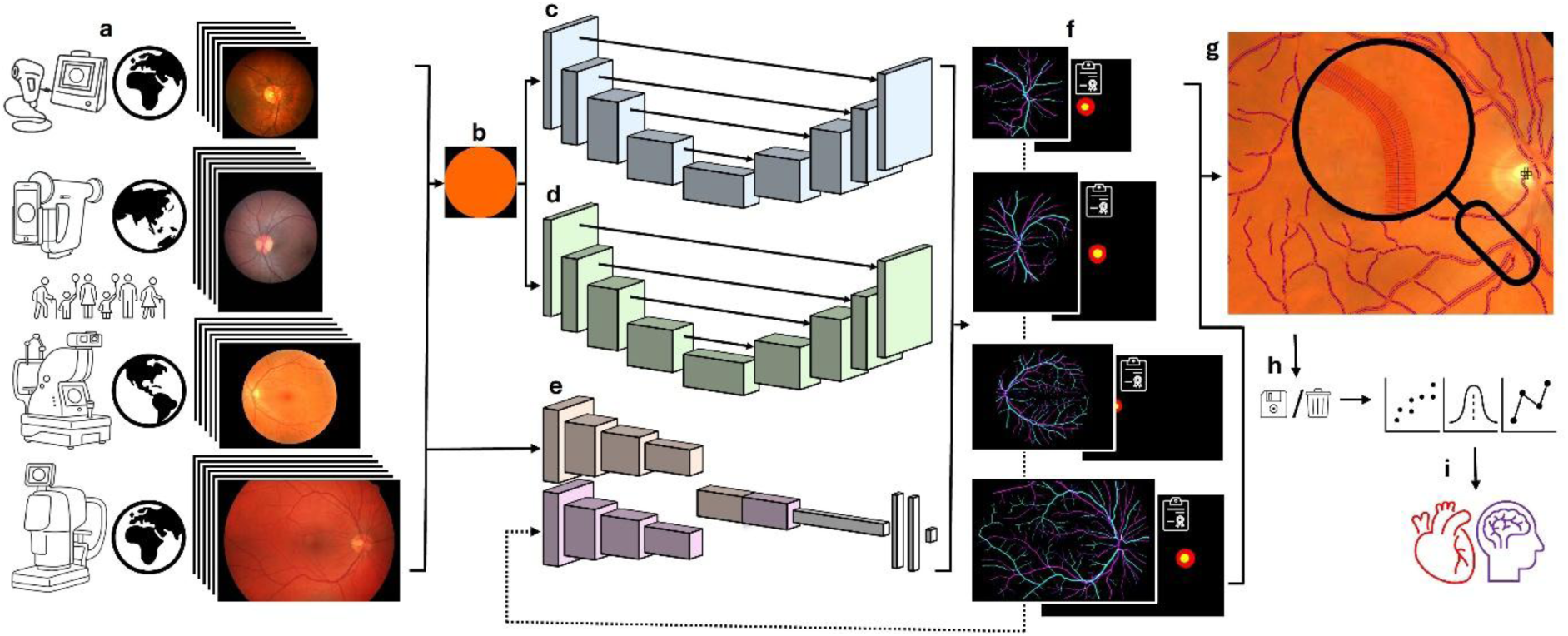
QUARTZ, from datasets to biomarker discovery. (a) Multiple, large, and varying retinal image datasets, (b) pre-processing, (c) arteriole/venule segmentation, (d) optic disc/cup segmentation, (e) image quality classification, (f) outputs, (g) vessel morphometric analysis, (h) data exclusion, and (i) statistical modelling for biomarker discovery.

High quality data annotation is crucial in training reliable deep learning models; therefore, the inclusion criteria for this study were strict. Inconsistencies were particularly evident in datasets annotated via crowdsourcing. For the datasets included in this study, any discrepancies in the annotations were corrected. Furthermore, since annotation styles can vary, annotations were adapted where necessary to conform to standardised rules.

### (3.1) Pre-processing

Each colour fundus image was normalised by cropping to the circular FOV and then adjusted to achieve a square shape by either centrally cropping or evenly zero padding the vertical dimension. Zero padding was important for images with the circular FOV truncated at the top and bottom.

### (3.2) Arteriole/Venule Segmentation

The U-Net architecture [47] was modified using ConvNeXt blocks [48] and was used for arteriole/venule (A/V) segmentation. The U-Net was further modified to employ a multi-segmentation technique [49] where the network outputs three independent channels to generate separate binary segmentation maps for the structures of arterioles, venules, and vessels. The model was pretrained via self-supervised learning using the 88,702 retinal images from the Kaggle EyePACS dataset [31].

All annotated images from the datasets were inspected and corrected by an expert human observer, limited to resolving annotation artefacts and colour inconsistencies. Pixels were labelled by colour, and any that did not match the defined class colours were corrected. In addition, single isolated labelled pixels and spurious labelled objects were removed.

For A/V segmentation, both four- and five-class [35] annotation schemes are common; the latter was adopted as it provides better distinction into more suitable categories. Where necessary, datasets were first standardised to the five-class approach, with the pixel labels of arteriole, venule, crossing (arteriole and venule overlap), uncertain (unclassifiable vessel), and background. Annotations were then converted to the desired model output format, with each channel represented as a binary map with the positive classes of arterioles, venules, and all vessels, respectively. Crossing pixels belonged to all three, and uncertain pixels belonged only to vessels.

Uncertain pixels were masked out of the binary maps for arterioles and venules. Each dataset was divided using a random 60:20:20 split for training, validation, and testing. However, if a dataset had a predefined split, that was used instead. For datasets with only a reported train and test split, a 75:25 split was applied to the training set to create a validation set. If images from the same participant were present, they were confined to a single set to prevent data leakage. The EPIC-Norfolk and Fundus-AVSeg annotated datasets were not split; the entire datasets were used exclusively as test sets.

The annotated datasets were of unequal size, containing between 22 and 240 images. To prevent the model from becoming biased toward larger datasets, a balanced sampling strategy was employed during training and validation so that each dataset contributed equally. Larger datasets were undersampled at each epoch to match the size of the UKBB set, while smaller datasets were oversampled to reach the same effective sample size. To ensure that no data were permanently excluded, the undersampling of larger datasets was performed dynamically. At the start of each epoch, the data were reshuffled and a different random subset was selected, allowing all available samples to be used over time.

The pre-processed images were resized to 1024 × 1024 pixels for model input (output was the same size). Augmentation of the training set included flips, scaling, translation, rotation, and adjustments to brightness, contrast, and saturation. Training used a batch size of 2 with the AdamW optimiser (learning rate = 0.001, weight decay = 0.005) for 1000 epochs. An exponential moving average (EMA) of the model weights with momentum = 0.99 was used. Additional learning rate decay showed no benefit. The loss function was a pixel-wise binary cross-entropy summed across channels; weighting the loss provided no improvement. The model was saved at the epoch with the lowest validation loss. The model was fit on the training set, with the validation set used for hyperparameter selection.

The output probability maps of size 1024 × 1024 pixels were returned to their original dimensions prior to pre- processing and then thresholded to produce binary segmentation maps. Rather than being optimised for a single dataset, the system required a single probability threshold applicable across all datasets. This was determined by selecting the threshold that maximised the average F1 score using the full validation sets (computed separately for each dataset and then averaged). Simple post-processing [50] was used to eliminate spurious objects.

Following the creation of vessel segments (see section 3.5), the scores from the arteriole and venule probability maps were used as soft votes and accumulated along the vessel segment centreline pixels to determine the A/V probability for each vessel segment.

### (3.3) Optic Disc/Cup Segmentation

The modified U-Net architecture in the A/V segmentation module was repurposed for optic disc/cup segmentation. The network outputs two independent channels to generate separate binary segmentation maps for the structures of optic disc and optic cup. Pretraining was performed using the 1200 annotated retinal images from the REFUGE dataset [42].

Some datasets only provided optic disc/cup contours and required converting into annotated images. All annotated images from the datasets were inspected and corrected by an expert human observer, with corrections limited to annotation artefacts and colour inconsistencies. All datasets followed a three-class scheme with pixel labels of disc, cup, and background; datasets following a two-class scheme (disc and background) were not collected. Annotations were then converted to the desired model output format, with each channel represented as a binary map with the positive classes of disc and cup, respectively. Importantly, since the cup region lies within the disc, pixels labelled as cup were also labelled as disc. The same dataset split portions (60:20:20) and rules from the A/V segmentation module were applied. However, the EPIC-Norfolk and ORIGA annotated datasets were not split; the entire datasets were reserved exclusively as test sets. The annotated datasets used for training and validation were of unequal size, ranging from 100 to 1345 images, so the same balanced sampling strategy as for the A/V segmentation module was applied.

The training approach was similar to that used for the A/V segmentation module, with the following adjustments: pre-processed images resized to 512 x 512 pixels, used a batch size of 8, trained for 500 epochs, with EMA of model weights removed, and employed a cosine decay learning rate schedule.

The output probability maps of size 512 × 512 pixels were returned to their original dimensions prior to pre- processing and then thresholded to produce binary segmentation maps. As with the A/V segmentation module, a single probability threshold applicable across all datasets was determined by selecting the threshold that maximised the average F1 score using the full validation sets. The largest segmented connected component determined the object of interest in each map. Optic disc localisation was determined from the centroid of the segmented disc. Additionally, the segmented disc and cup were used to calculate the vertical cup-to-disc ratio (vCDR) [42].

### (3.4) Image Quality Classification

Large retinal datasets used in cohort studies often contain a number of lower quality images, particularly those with partial illumination caused by inadequate pupil dilation or inherently small pupils. Additional quality issues include blur and obstruction resulting from focal plane errors, eyelashes or blinking, lens artefacts, media opacities (such as cataracts), poor head position, and head or eye movement. Nevertheless, valuable information can still be extracted from accurately segmented portions of the vasculature, even if this only represents a section of the vascular tree. A dual model based on concatenating outputs from two instances of the EfficientNetV2-S architecture [51] was developed for image quality classification for epidemiological studies. One network took A/V segmentation maps as input, while the other took colour retinal images as input. Each EfficientNetV2-S instance was initially pretrained on the ImageNet dataset, followed by the entire model being further pretrained using 28,792 labelled retinal images from the EyeQ dataset and their output from the A/V segmentation module.

The non-public datasets used in this module inherently conformed to annotation standards, whereas the FQS dataset had to be relabelled by an expert human observer. Each dataset followed a two-class scheme, with image labels of either inadequate or adequate based on the usability of the A/V segmented maps in epidemiological studies. The inadequate label included non-retinal images, which are present in all real-world large datasets. Full labelling criteria have been described previously [12]. The proportion of adequate quality images was 76.65% in UKBB, 84.10% in CLSA, 87.41% in NEL DESP (64.90% when including non-retinal images), 81.52% in FQS, and 72.75% in EPIC-Norfolk. Each dataset was randomly split into 80:10:10 training, validation, and test sets. However, the EPIC-Norfolk and FQS annotated datasets were not split; the entire datasets were used exclusively as test sets. As all annotated datasets used for training and validation were of equal size, no balanced sampling strategy was required.

Pre-processing was avoided because the FOV could not always be reliably identified in some of the very lowest quality images, and retaining its full context within the frame was also important. Images were resized to 384 × 384 pixels for model input. Data augmentation was consistent with other modules, except that translation and scaling were excluded, as these could alter the proportion of retina captured, an important indicator of image quality. Training used a batch size of 16 with the AdamW optimiser (learning rate = 0.001, weight decay = 0.005), additional learning rate decay showed no benefit. The three-class classification layer of the model pretrained on EyeQ was replaced with a sigmoid classification layer with randomly initialised weights. The model was initially trained for 5 epochs for the classification layer, followed by 20 epochs for the rest of the top layers. The loss function was binary cross-entropy; weighting the loss offered no improvements. The model was saved at the epoch with the lowest validation loss. The model was fit on the training set, with the validation set used for hyperparameter selection. As with the segmentation modules, a single probability threshold applicable across all datasets was determined by selecting the threshold that maximised the average F1 score using the validation sets.

### (3.5) Vessel Morphometric Analysis

Morphological thinning was applied to the vessel binary map from the A/V segmentation module to create centrelines, followed by removal of spurs, branching, and crossover points to create vessel segments, which were then smoothed using parametric spline curve fitting [50]. Analysis of the vessel segments, which included measurements of width and tortuosity, used previously well documented and validated techniques with previously demonstrated cross-dataset robustness [39,50].

Whilst the deep learning modules mitigate image size variability through cropping and resizing, enabling consistent feature extraction across datasets of varying resolutions. Conversely, the morphometric vessel module was performed on the original image dimensions and included several size dependent variables. Following our previous studies [12,39,50], these variables have been adjusted by scaling according to the horizontal FOV diameter to account for differences in image size. It is important to note that this approach assumes a consistent FOV angle across all images in a dataset, which is already standard practice in most study designs.

### (3.6) QUARTZ Online Tool

To support the research community, the generalisation-focused version of QUARTZ has been released as an online tool (https://miqa.kingston.ac.uk/quartz/), providing broad accessibility and enabling reproducible biomarker research across a wide range of datasets. The online version offers a simple interface where images can be easily dragged and dropped, with segmentations and full morphometric measures available for download. Comprehensive documentation is available online and includes recommended usage instructions, detailed descriptions of outputs with a complete list of measurements, and guidance on the use of these outputs.

QUARTZ has been trained to reliably analyse colour retinal images with a FOV between 30° and 60°, which reflects the most common settings used in retinal imaging. However, vessel morphometric measures are not directly comparable across images with differing FOV angles. Therefore, when using QUARTZ, it is important to maintain a consistent FOV angle within a study, which is typically ensured by the imaging protocol.

Variations in FOV angle can alter vessel distribution and affect the FOV-based size adjustments described in Section 3.5. QUARTZ is robust to typical retinal pathology and variations in illumination, but extreme pathology or non-standard imaging protocols (e.g., images with atypical filters) may reduce the accuracy of vessel segmentations and morphometric measures. Whilst images with inadequate segmentation will be excluded based on quality, users are also able to review visually the segmentations to support responsible use and reproducible adoption of the tool.

### (3.7) Camera-Level Repeatability

To gain insight into the reliability of QUARTZ using existing data, repeatability across multiple imaging devices was assessed using images from a single participant whose retina was photographed on the same day using 32 cameras across 15 locations within the NEL DESP screening programme. This analysis specifically explored camera-related variation in vessel width measurements. Camera models included Canon CR2 AF, Topcon NW6, and OCT Topcon Maestro. Only macula-centred images of the right eye were used for this assessment.

For comparison, 100 participants were randomly selected from the NEL DESP dataset [30], with the macula-centred image of the right eye used for each. Ideally, all participants would have been imaged on a single camera; however, camera metadata were unavailable for this dataset. Consequently, the observed variation reflects both biological variation between participants and camera-related variation.

The images of various resolutions were processed using QUARTZ, which outputs vessel measurements in pixels that were subsequently converted to microns using a conversion factor derived from the vertical diameter of the optic disc [52]. To ensure high certainty in vessel type identification, segments with an A/V probability < 0.8 were excluded. Analyses focused on larger retinal vessels, with vessel segments having a mean diameter < 50 microns being excluded. Vessel widths were then summarised as image-level means (contribution of each vessel segment was weighted by its segment length), consistent with previous epidemiological studies using QUARTZ outputs [3,18,25].

## (4) Results

Evaluations were conducted on the test sets. In this section, the suffix -GEN is appended to QUARTZ to denote the new generic version, which is generalisation-focused, while -DL refers to the earlier version that transitioned to a deep learning pipeline and was tuned for specific datasets. Reported performance metrics include sensitivity, specificity, precision (positive predictive value), negative predictive value (NPV), accuracy, F1 score, intersection over union (IoU), area under the receiver operating characteristic curve (AUC ROC), and area under the precision-recall curve (AUC PR).

Table 2 reports the A/V segmentation results, with binary segmentations evaluated separately for arterioles, venules, and vessels. The evaluation used only pixels within the circular FOV. Pixels annotated as uncertain were excluded for arteriole and venule assessments but retained for vessel evaluation. Across the nine internal datasets, QUARTZ-GEN demonstrated consistently high performance across all vessel types. For arteriole segmentation, F1 scores ranged from 0.7559 to 0.8306, with a mean F1 of 0.7956 ± 0.0236 and a coefficient of variation (CV) of 2.97%. For venule segmentation, F1 scores ranged from 0.7894 to 0.8452, with a mean F1 of 0.8200 ± 0.0197 and a CV of 2.40%. For vessel segmentation, F1 scores ranged from 0.8020 to 0.8698, with a mean F1 of 0.8475 ± 0.0179 and a CV of 2.12%. This indicates a low variability and stable performance across the different datasets. On the external EPIC-Norfolk dataset, QUARTZ-GEN achieved F1 scores of 0.8477 for venules and 0.8741 for vessels, slightly exceeding previously observed maxima, while attaining near-peak performance for arterioles (0.8305). See Figure 2 for examples of A/V segmentation outputs across different datasets, and Figure 4 for a visual comparison of F1 performance.

**Figure 2:**
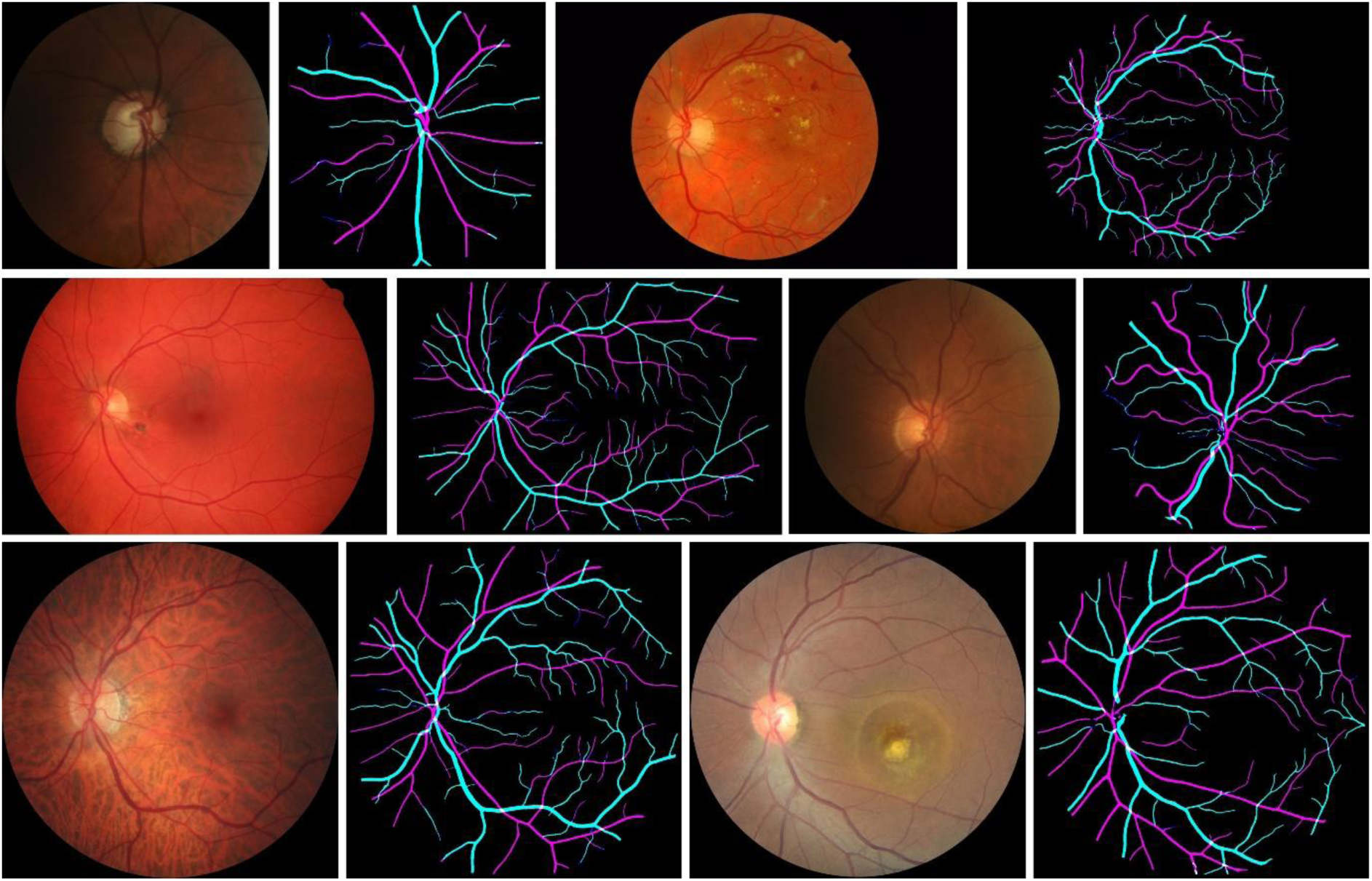
QUARTZ-GEN A/V segmentation examples on retinal images from the Leuven-Haifa, MESSIDOR [53], HRF-AV, LES-AV, and Fundus-AVSeg datasets (two examples shown for Fundus-AVSeg), displayed left-to-right, top-to-bottom. The segmentations depict the RGB composition of three independent binary segmentation maps for the structures of arterioles, venules, and vessels.

**Figure 3:**
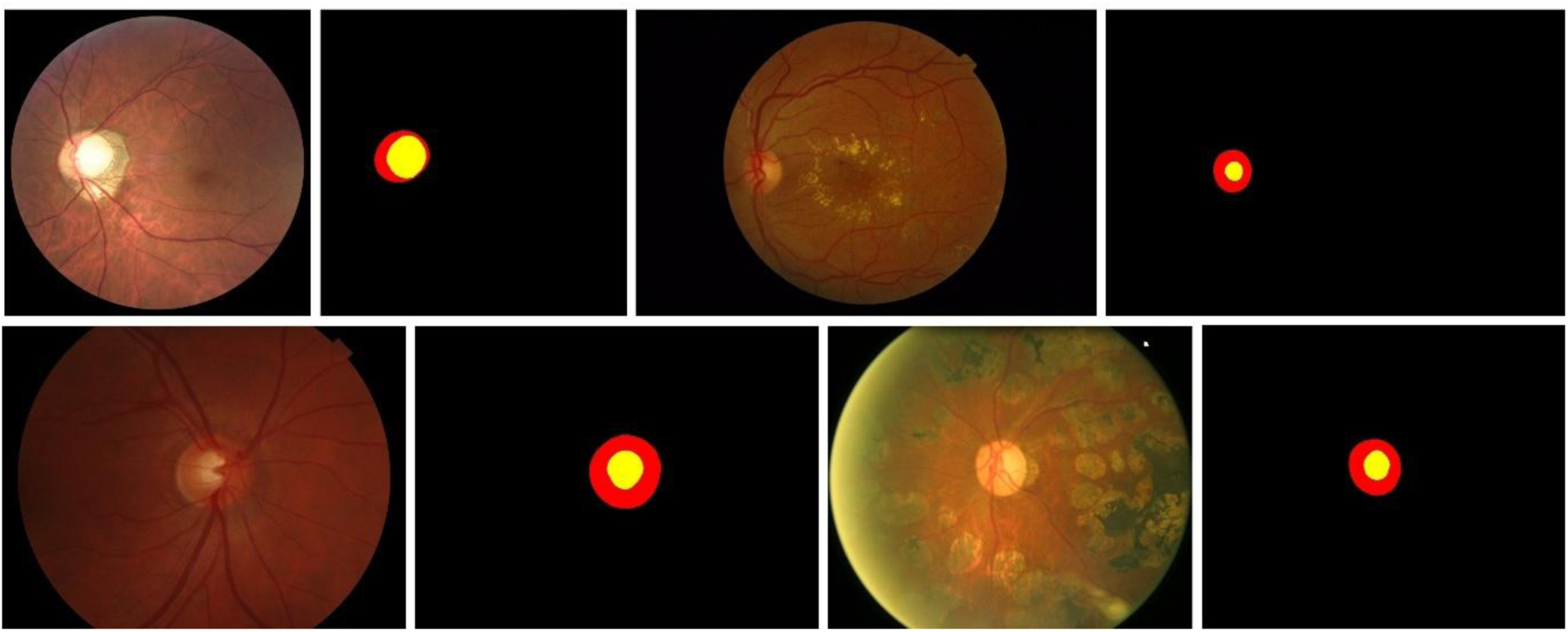
QUARTZ-GEN optic disc/cup segmentation examples on retinal images from the REFUGE, MESSIDOR [53], PAPILA, and ORIGA datasets, displayed left-to-right, top-to-bottom. The segmentations depict the RGB composition of two independent binary segmentation maps for the structures of the disc and cup, with the third channel left empty.

**Figure 4:**
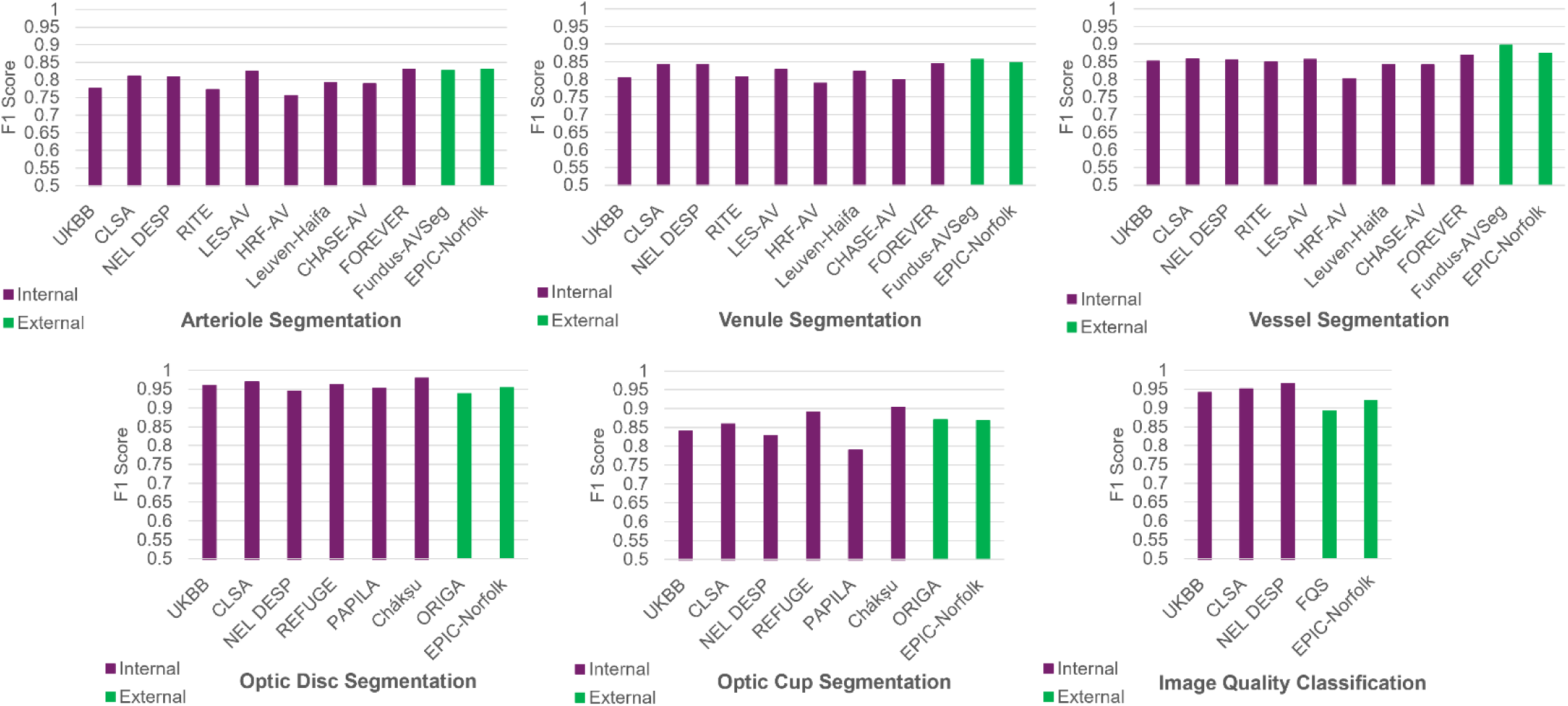
QUARTZ-GEN cross-dataset performance on internal and external datasets, shown for the segmentation and classification tasks.

**Table 2:** Performance of A/V segmentation (prior to post-processing). (i) denotes internal dataset; (e) denotes external dataset.

| Method | Dataset | Map | Sensitivity | Specificity | Precision | Accuracy | F<sub>1</sub> Score | IoU | AUC ROC | AUC PR |
| --- | --- | --- | --- | --- | --- | --- | --- | --- | --- | --- |
| QUARTZ-DL | UKBB (i) | Arteriole | 0.7699 | 0.9875 | 0.7526 | 0.9773 | 0.7612 | 0.6144 | 0.9866 | 0.8479 |
|  |  | Venule | 0.7832 | 0.9902 | 0.8144 | 0.9794 | 0.7985 | 0.6645 | 0.9901 | 0.8895 |
|  |  | Vessel | 0.8545 | 0.9814 | 0.8401 | 0.9684 | 0.8472 | 0.7349 | 0.9896 | 0.9313 |
| QUARTZ-DL | CLSA (i) | Arteriole | 0.7757 | 0.9872 | 0.7642 | 0.9764 | 0.7699 | 0.6259 | 0.9859 | 0.8499 |
|  |  | Venule | 0.7976 | 0.9894 | 0.8187 | 0.9785 | 0.8080 | 0.6779 | 0.9893 | 0.8961 |
|  |  | Vessel | 0.8581 | 0.9803 | 0.8445 | 0.9667 | 0.8513 | 0.7410 | 0.9883 | 0.9312 |
| QUARTZ-DL | NEL DESP (i) | Arteriole | 0.7627 | 0.9905 | 0.7861 | 0.9806 | 0.7742 | 0.6316 | 0.9878 | 0.8585 |
|  |  | Venule | 0.8046 | 0.9913 | 0.8262 | 0.9821 | 0.8153 | 0.6881 | 0.9908 | 0.8966 |
|  |  | Vessel | 0.8462 | 0.9836 | 0.8471 | 0.9703 | 0.8467 | 0.7341 | 0.9896 | 0.9309 |
| QUARTZ-GEN | UKBB (i) | Arteriole | 0.7893 | 0.9878 | 0.7623 | 0.9785 | 0.7756 | 0.6334 | 0.9891 | 0.8506 |
|  |  | Venule | 0.8115 | 0.9886 | 0.7966 | 0.9794 | 0.8040 | 0.6722 | 0.9897 | 0.8939 |
|  |  | Vessel | 0.8765 | 0.9793 | 0.8286 | 0.9687 | 0.8519 | 0.7420 | 0.9908 | 0.9355 |
| QUARTZ-GEN | CLSA (i) | Arteriole | 0.8067 | 0.9902 | 0.8155 | 0.9809 | 0.8111 | 0.6822 | 0.9921 | 0.8903 |
|  |  | Venule | 0.8376 | 0.9909 | 0.8466 | 0.9822 | 0.8420 | 0.7272 | 0.9929 | 0.9253 |
|  |  | Vessel | 0.8570 | 0.9825 | 0.8591 | 0.9685 | 0.8581 | 0.7514 | 0.9908 | 0.9394 |
| QUARTZ-GEN | NEL DESP (i) | Arteriole | 0.7961 | 0.9921 | 0.8215 | 0.9835 | 0.8086 | 0.6787 | 0.9923 | 0.8898 |
|  |  | Venule | 0.8281 | 0.9929 | 0.8576 | 0.9848 | 0.8426 | 0.7280 | 0.9934 | 0.9224 |
|  |  | Vessel | 0.8477 | 0.9854 | 0.8617 | 0.9721 | 0.8547 | 0.7462 | 0.9911 | 0.9386 |
| QUARTZ-GEN | RITE (i) | Arteriole | 0.7554 | 0.9879 | 0.7909 | 0.9746 | 0.7727 | 0.6296 | 0.9804 | 0.8490 |
|  |  | Venule | 0.7868 | 0.9881 | 0.8289 | 0.9743 | 0.8073 | 0.6769 | 0.9830 | 0.8858 |
|  |  | Vessel | 0.8517 | 0.9784 | 0.8469 | 0.9628 | 0.8493 | 0.7381 | 0.9852 | 0.9306 |
| QUARTZ-GEN | LES-AV (i) | Arteriole | 0.8546 | 0.9899 | 0.7965 | 0.9840 | 0.8245 | 0.7015 | 0.9911 | 0.9026 |
|  |  | Venule | 0.8643 | 0.9892 | 0.7942 | 0.9834 | 0.8278 | 0.7062 | 0.9906 | 0.9138 |
|  |  | Vessel | 0.9031 | 0.9795 | 0.8150 | 0.9725 | 0.8568 | 0.7494 | 0.9905 | 0.9397 |
| QUARTZ-GEN | HRF-AV (i) | Arteriole | 0.7520 | 0.9900 | 0.7598 | 0.9804 | 0.7559 | 0.6076 | 0.9881 | 0.8345 |
|  |  | Venule | 0.7706 | 0.9909 | 0.8091 | 0.9804 | 0.7894 | 0.6520 | 0.9882 | 0.8767 |
|  |  | Vessel | 0.8085 | 0.9800 | 0.7957 | 0.9649 | 0.8020 | 0.6695 | 0.9849 | 0.8881 |
| QUARTZ-GEN | Leuven-Haifa (i) | Arteriole | 0.8225 | 0.9878 | 0.7649 | 0.9802 | 0.7927 | 0.6565 | 0.9885 | 0.8797 |
|  |  | Venule | 0.8178 | 0.9902 | 0.8277 | 0.9809 | 0.8227 | 0.6988 | 0.9884 | 0.9053 |
|  |  | Vessel | 0.8748 | 0.9780 | 0.8127 | 0.9679 | 0.8426 | 0.7280 | 0.9882 | 0.9285 |
| QUARTZ-GEN | CHASE-AV (i) | Arteriole | 0.8081 | 0.9863 | 0.7703 | 0.9768 | 0.7888 | 0.6512 | 0.9883 | 0.8756 |
|  |  | Venule | 0.7927 | 0.9892 | 0.8051 | 0.9787 | 0.7989 | 0.6651 | 0.9887 | 0.8826 |
|  |  | Vessel | 0.8583 | 0.9779 | 0.8262 | 0.9649 | 0.8419 | 0.7270 | 0.9878 | 0.9252 |
| QUARTZ-GEN | FOREVER (i) | Arteriole | 0.8189 | 0.9931 | 0.8426 | 0.9856 | 0.8306 | 0.7103 | 0.9942 | 0.9152 |
|  |  | Venule | 0.8641 | 0.9913 | 0.8271 | 0.9855 | 0.8452 | 0.7319 | 0.9941 | 0.9280 |
|  |  | Vessel | 0.8886 | 0.9846 | 0.8519 | 0.9759 | 0.8698 | 0.7696 | 0.9940 | 0.9496 |
| QUARTZ-GEN | Fundus-AVSeg (e) | Arteriole | 0.7956 | 0.9924 | 0.8615 | 0.9813 | 0.8272 | 0.7053 | 0.9883 | 0.8941 |
|  |  | Venule | 0.8138 | 0.9939 | 0.9057 | 0.9817 | 0.8573 | 0.7502 | 0.9887 | 0.9248 |
|  |  | Vessel | 0.8736 | 0.9898 | 0.9228 | 0.9755 | 0.8976 | 0.8141 | 0.9929 | 0.9635 |
| QUARTZ-GEN | EPIC-Norfolk (e) | Arteriole | 0.8067 | 0.9926 | 0.8558 | 0.9830 | 0.8305 | 0.7101 | 0.9932 | 0.9116 |
|  |  | Venule | 0.8240 | 0.9928 | 0.8729 | 0.9832 | 0.8477 | 0.7357 | 0.9930 | 0.9300 |
|  |  | Vessel | 0.8596 | 0.9867 | 0.8890 | 0.9727 | 0.8741 | 0.7763 | 0.9932 | 0.9543 |

**Table 3:** Performance of A/V segment-level decisions. (i) denotes internal dataset; (e) denotes external dataset.

| Method | Dataset | Class | Sensitivity | Specificity | Precision | Accuracy | F<sub>1</sub> Score |
| --- | --- | --- | --- | --- | --- | --- | --- |
| QUARTZ-DL | UKBB (i) | Arteriole | 0.9079 | 0.8971 | 0.8857 | 0.9022 | 0.8967 |
|  |  | Venule | 0.8971 | 0.9079 | 0.9173 | 0.9022 | 0.9071 |
| QUARTZ-DL | CLSA (i) | Arteriole | 0.8804 | 0.8713 | 0.8567 | 0.8755 | 0.8684 |
|  |  | Venule | 0.8713 | 0.8804 | 0.8929 | 0.8755 | 0.8820 |
| QUARTZ-DL | NEL DESP (i) | Arteriole | 0.8882 | 0.9290 | 0.9073 | 0.9111 | 0.8977 |
|  |  | Venule | 0.9290 | 0.8882 | 0.9140 | 0.9111 | 0.9214 |
| QUARTZ-GEN | UKBB (i) | Arteriole | 0.8986 | 0.9106 | 0.9018 | 0.9048 | 0.9002 |
|  |  | Venule | 0.9106 | 0.8986 | 0.9076 | 0.9048 | 0.9091 |
| QUARTZ-GEN | CLSA (i) | Arteriole | 0.9644 | 0.9737 | 0.9683 | 0.9695 | 0.9663 |
|  |  | Venule | 0.9737 | 0.9644 | 0.9705 | 0.9695 | 0.9721 |
| QUARTZ-GEN | NEL DESP (i) | Arteriole | 0.9289 | 0.9392 | 0.9202 | 0.9348 | 0.9245 |
|  |  | Venule | 0.9392 | 0.9289 | 0.9459 | 0.9348 | 0.9425 |
| QUARTZ-GEN | RITE (i) | Arteriole | 0.9467 | 0.9565 | 0.9480 | 0.9520 | 0.9474 |
|  |  | Venule | 0.9565 | 0.9467 | 0.9554 | 0.9520 | 0.9560 |
| QUARTZ-GEN | LES-AV (i) | Arteriole | 0.9406 | 0.9661 | 0.9596 | 0.9543 | 0.9500 |
|  |  | Venule | 0.9661 | 0.9406 | 0.9500 | 0.9543 | 0.9580 |
| QUARTZ-GEN | HRF-AV (i) | Arteriole | 0.9716 | 0.9359 | 0.9292 | 0.9525 | 0.9500 |
|  |  | Venule | 0.9359 | 0.9716 | 0.9744 | 0.9525 | 0.9548 |
| QUARTZ-GEN | Leuven-Haifa (i) | Arteriole | 0.9529 | 0.9498 | 0.9496 | 0.9513 | 0.9513 |
|  |  | Venule | 0.9498 | 0.9529 | 0.9531 | 0.9513 | 0.9514 |
| QUARTZ-GEN | CHASE-AV (i) | Arteriole | 0.9341 | 0.9231 | 0.9231 | 0.9286 | 0.9286 |
|  |  | Venule | 0.9231 | 0.9341 | 0.9341 | 0.9286 | 0.9286 |
| QUARTZ-GEN | FOREVER (i) | Arteriole | 0.9135 | 0.9672 | 0.9548 | 0.9441 | 0.9337 |
|  |  | Venule | 0.9672 | 0.9135 | 0.9365 | 0.9441 | 0.9516 |
| QUARTZ-GEN | Fundus-AVSeg (e) | Arteriole | 0.9218 | 0.9362 | 0.9223 | 0.9297 | 0.9220 |
|  |  | Venule | 0.9362 | 0.9218 | 0.9358 | 0.9297 | 0.9360 |
| QUARTZ-GEN | EPIC-Norfolk (e) | Arteriole | 0.9433 | 0.9503 | 0.9426 | 0.9471 | 0.9429 |
|  |  | Venule | 0.9503 | 0.9433 | 0.9509 | 0.9471 | 0.9506 |

When comparing QUARTZ-GEN to QUARTZ-DL, A/V segmentation performance improvements were observed across all vessel types. For UKBB, arteriole F1 increased from 0.7612 to 0.7756, venule F1 increased from 0.7985 to 0.8040, and vessel F1 increased from 0.8472 to 0.8519. Larger improvements were observed for the CLSA and NEL DESP datasets. Figure 5 shows a visual comparison of F1 performance. QUARTZ-GEN generally achieved higher sensitivities while maintaining comparable specificities.

**Figure 5:**
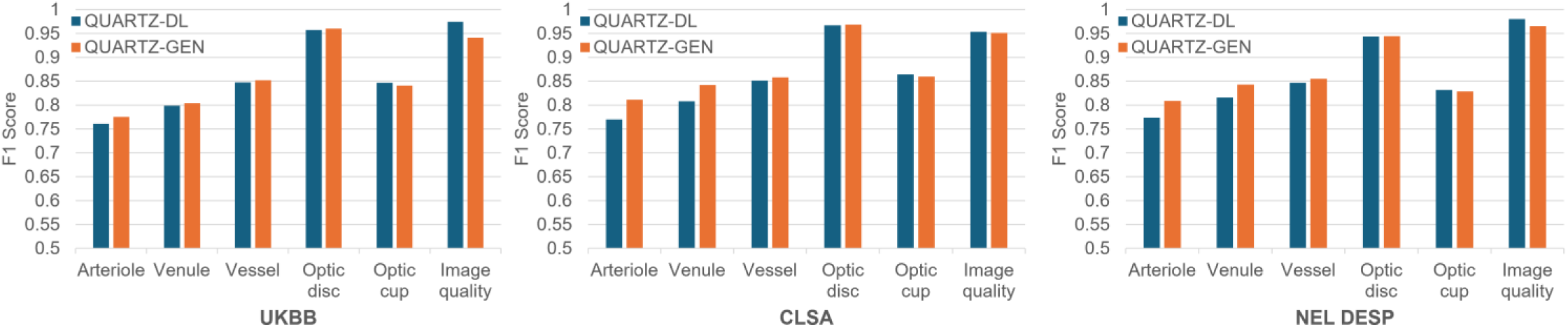
Performance comparison between QUARTZ-DL and QUARTZ-GEN across UKBB, CLSA and NEL DESP datasets, shown for the segmentation and classification tasks.

Tables 3 summarises the A/V performance on a vessel segment-level basis. QUARTZ-GEN demonstrated consistently high performance and low variability across the nine internal datasets. The F1 scores ranged from 0.9002 to 0.9663 for arterioles, with a mean F1 of 0.9391 ± 0.0184 and a CV of 1.96%. The F1 score ranged from 0.9091 to 0.9721 for venules, with a mean F1 of 0.9471 ± 0.0174 and a CV of 1.84%. On the external EPIC-Norfolk dataset, QUARTZ-GEN maintained a high performance, achieving F1 scores of 0.9429 for arterioles and 0.9506 for venules. Comparing the transition from QUARTZ-DL to QUARTZ-GEN, for UKBB, the F1 scores showed modest improvements, increasing from 0.8967 to 0.9002 for arteriole and from 0.9071 to 0.9091 for venules. The CLSA and NEL DESP datasets exhibited more pronounced gains. To further increase certainty of correct vessel type identification, epidemiological studies [3] have adopted a 0.8 probability threshold at the cost of retaining only a percentage of the vessel segments as shown in Table 4.

**Table 4:** Performance of A/V segment-level decision with an arteriole > 0.8 and venule > 0.8 probability threshold applied. (i) denotes internal dataset; (e) denotes external dataset.

| Method | Dataset | Class | Sensitivity | Specificity | Precision | Accuracy | F<sub>1</sub> Score | Fraction of Data Retained |
| --- | --- | --- | --- | --- | --- | --- | --- | --- |
| QUARTZ-DL | UKBB (i) | Arteriole | 0.9941 | 0.9709 | 0.9657 | 0.9814 | 0.9797 | 0.7358 |
|  |  | Venule | 0.9709 | 0.9941 | 0.9950 | 0.9814 | 0.9828 |  |
| QUARTZ-DL | CLSA (i) | Arteriole | 0.9655 | 0.9603 | 0.9532 | 0.9627 | 0.9593 | 0.7282 |
|  |  | Venule | 0.9603 | 0.9655 | 0.9708 | 0.9627 | 0.9655 |  |
| QUARTZ-DL | NEL DESP (i) | Arteriole | 0.9418 | 0.9774 | 0.9687 | 0.9622 | 0.9551 | 0.7843 |
|  |  | Venule | 0.9774 | 0.9418 | 0.9577 | 0.9622 | 0.9674 |  |
| QUARTZ-GEN | UKBB (i) | Arteriole | 0.9654 | 0.9542 | 0.9489 | 0.9594 | 0.9571 | 0.8529 |
|  |  | Venule | 0.9542 | 0.9654 | 0.9690 | 0.9544 | 0.9615 |  |
| QUARTZ-GEN | CLSA (i) | Arteriole | 0.9785 | 0.9825 | 0.9785 | 0.9807 | 0.9785 | 0.9318 |
|  |  | Venule | 0.9825 | 0.9785 | 0.9825 | 0.9807 | 0.9825 |  |
| QUARTZ-GEN | NEL DESP (i) | Arteriole | 0.9740 | 0.9702 | 0.9614 | 0.9718 | 0.9677 | 0.9052 |
|  |  | Venule | 0.9702 | 0.9740 | 0.9800 | 0.9718 | 0.9751 |  |
| QUARTZ-GEN | RITE (i) | Arteriole | 0.9710 | 0.9746 | 0.9696 | 0.9730 | 0.9703 | 0.9220 |
|  |  | Venule | 0.9746 | 0.9710 | 0.9758 | 0.9730 | 0.9752 |  |
| QUARTZ-GEN | LES-AV (i) | Arteriole | 0.9783 | 0.9910 | 0.9890 | 0.9852 | 0.9836 | 0.9269 |
|  |  | Venule | 0.9910 | 0.9783 | 0.9821 | 0.9852 | 0.9865 |  |
| QUARTZ-GEN | HRF-AV (i) | Arteriole | 0.9906 | 0.9805 | 0.9787 | 0.9853 | 0.9846 | 0.8947 |
|  |  | Venule | 0.9805 | 0.9906 | 0.9914 | 0.9853 | 0.9859 |  |
| QUARTZ-GEN | Leuven-Haifa (i) | Arteriole | 0.9744 | 0.9807 | 0.9810 | 0.9775 | 0.9777 | 0.9044 |
|  |  | Venule | 0.9807 | 0.9744 | 0.9739 | 0.9775 | 0.9773 |  |
| QUARTZ-GEN | CHASE-AV (i) | Arteriole | 0.9735 | 0.9595 | 0.9608 | 0.9666 | 0.9671 | 0.8899 |
|  |  | Venule | 0.9595 | 0.9735 | 0.9726 | 0.9666 | 0.9660 |  |
| QUARTZ-GEN | FOREVER (i) | Arteriole | 0.9750 | 0.9870 | 0.9811 | 0.9821 | 0.9781 | 0.9114 |
|  |  | Venule | 0.9870 | 0.9750 | 0.9828 | 0.9821 | 0.9849 |  |
| QUARTZ-GEN | Fundus-AVSeg (e) | Arteriole | 0.9507 | 0.9583 | 0.9485 | 0.9549 | 0.9496 | 0.9157 |
|  |  | Venule | 0.9583 | 0.9507 | 0.9600 | 0.9549 | 0.9591 |  |
| QUARTZ-GEN | EPIC-Norfolk (e) | Arteriole | 0.9810 | 0.9736 | 0.9692 | 0.9770 | 0.9751 | 0.9087 |
|  |  | Venule | 0.9736 | 0.9810 | 0.9837 | 0.9770 | 0.9786 |  |

The results of optic disc/cup segmentation are reported in Table 5, with binary segmentations evaluated separately for the disc and cup. The evaluation used only pixels within the circular FOV. Across the six internal datasets, QUARTZ-GEN demonstrated consistently high performance for disc segmentation and generally high performance for cup segmentation. For disc segmentation, F1 scores ranged from 0.9444 to 0.9782, with a mean F1 of 0.9611 ± 0.0108 and a CV of 1.13%, indicating very low variability across datasets. For cup segmentation, F1 scores ranged from 0.7893 to 0.9044, with a mean F1 of 0.8520 ± 0.0385 and a CV of 4.52%, indicating low-to-moderate variability, with the PAPILA dataset contributing most to this variation. On the external EPIC-Norfolk dataset, QUARTZ-GEN maintained a high performance, achieving F1 scores of 0.9532 for disc and 0.8677 for cup. Examples of outputs from optic disc/cup segmentation across different datasets are provided in Figure 3 and a visual comparison of F1 performance is provided in Figure 4.

**Table 5:**
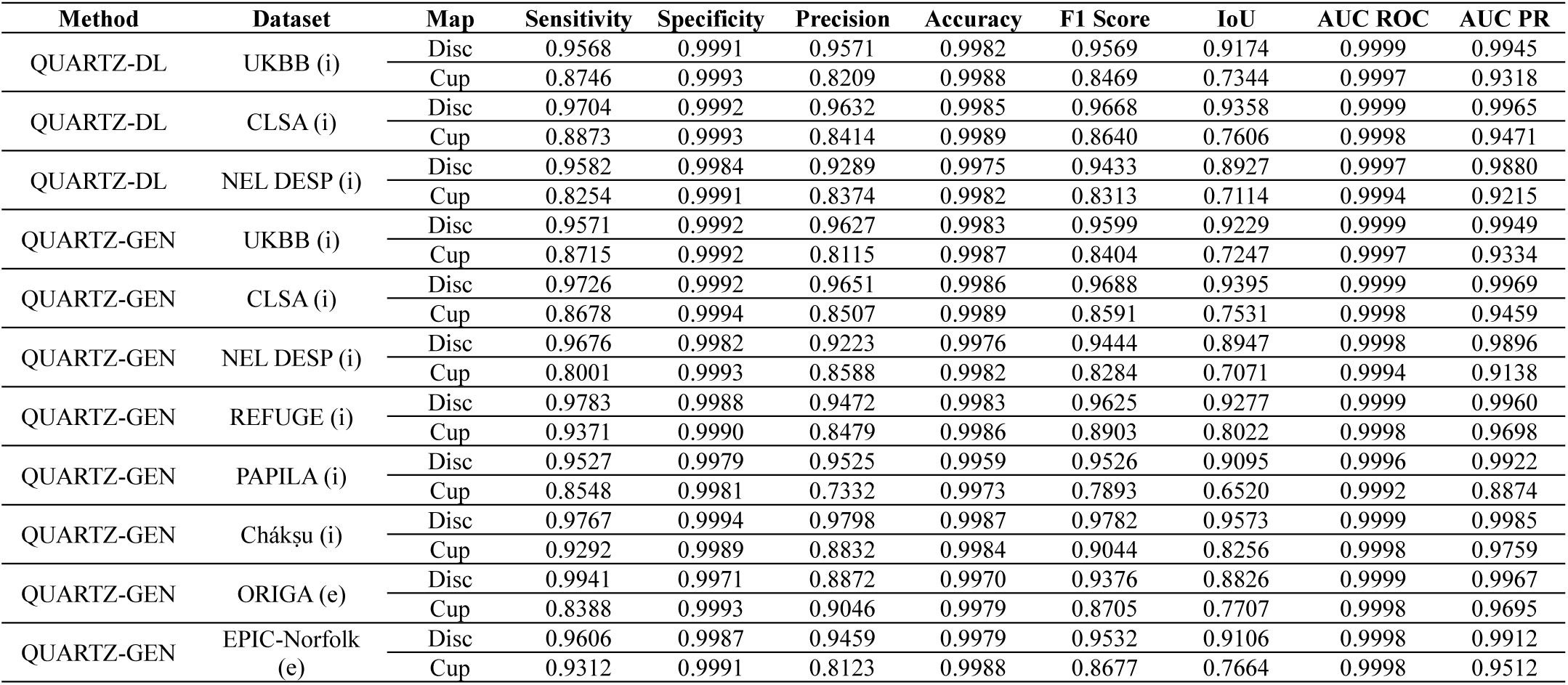
Performance of optic disc/cup segmentation. (i) denotes internal dataset; (e) denotes external dataset.

| Method | Dataset | Map | Sensitivity | Specificity | Precision | Accuracy | F1 Score | IoU | AUC ROC | AUC PR |
| --- | --- | --- | --- | --- | --- | --- | --- | --- | --- | --- |
| QUARTZ-DL | UKBB (i) | Disc | 0.9568 | 0.9991 | 0.9571 | 0.9982 | 0.9569 | 0.9174 | 0.9999 | 0.9945 |
|  |  | Cup | 0.8746 | 0.9993 | 0.8209 | 0.9988 | 0.8469 | 0.7344 | 0.9997 | 0.9318 |
| QUARTZ-DL | CLSA (i) | Disc | 0.9704 | 0.9992 | 0.9632 | 0.9985 | 0.9668 | 0.9358 | 0.9999 | 0.9965 |
|  |  | Cup | 0.8873 | 0.9993 | 0.8414 | 0.9989 | 0.8640 | 0.7606 | 0.9998 | 0.9471 |
| QUARTZ-DL | NEL DESP (i) | Disc | 0.9582 | 0.9984 | 0.9289 | 0.9975 | 0.9433 | 0.8927 | 0.9997 | 0.9880 |
|  |  | Cup | 0.8254 | 0.9991 | 0.8374 | 0.9982 | 0.8313 | 0.7114 | 0.9994 | 0.9215 |
| QUARTZ-GEN | UKBB (i) | Disc | 0.9571 | 0.9992 | 0.9627 | 0.9983 | 0.9599 | 0.9229 | 0.9999 | 0.9949 |
|  |  | Cup | 0.8715 | 0.9992 | 0.8115 | 0.9987 | 0.8404 | 0.7247 | 0.9997 | 0.9334 |
| QUARTZ-GEN | CLSA (i) | Disc | 0.9726 | 0.9992 | 0.9651 | 0.9986 | 0.9688 | 0.9395 | 0.9999 | 0.9969 |
|  |  | Cup | 0.8678 | 0.9994 | 0.8507 | 0.9989 | 0.8591 | 0.7531 | 0.9998 | 0.9459 |
| QUARTZ-GEN | NEL DESP (i) | Disc | 0.9676 | 0.9982 | 0.9223 | 0.9976 | 0.9444 | 0.8947 | 0.9998 | 0.9896 |
|  |  | Cup | 0.8001 | 0.9993 | 0.8588 | 0.9982 | 0.8284 | 0.7071 | 0.9994 | 0.9138 |
| QUARTZ-GEN | REFUGE (i) | Disc | 0.9783 | 0.9988 | 0.9472 | 0.9983 | 0.9625 | 0.9277 | 0.9999 | 0.9960 |
|  |  | Cup | 0.9371 | 0.9990 | 0.8479 | 0.9986 | 0.8903 | 0.8022 | 0.9998 | 0.9698 |
| QUARTZ-GEN | PAPILA (i) | Disc | 0.9527 | 0.9979 | 0.9525 | 0.9959 | 0.9526 | 0.9095 | 0.9996 | 0.9922 |
|  |  | Cup | 0.8548 | 0.9981 | 0.7332 | 0.9973 | 0.7893 | 0.6520 | 0.9992 | 0.8874 |
| QUARTZ-GEN | Chákşu (i) | Disc | 0.9767 | 0.9994 | 0.9798 | 0.9987 | 0.9782 | 0.9573 | 0.9999 | 0.9985 |
|  |  | Cup | 0.9292 | 0.9989 | 0.8832 | 0.9984 | 0.9044 | 0.8256 | 0.9998 | 0.9759 |
| QUARTZ-GEN | ORIGA (e) | Disc | 0.9941 | 0.9971 | 0.8872 | 0.9970 | 0.9376 | 0.8826 | 0.9999 | 0.9967 |
|  |  | Cup | 0.8388 | 0.9993 | 0.9046 | 0.9979 | 0.8705 | 0.7707 | 0.9998 | 0.9695 |
| QUARTZ-GEN | EPIC-Norfolk (e) | Disc | 0.9606 | 0.9987 | 0.9459 | 0.9979 | 0.9532 | 0.9106 | 0.9998 | 0.9912 |
|  |  | Cup | 0.9312 | 0.9991 | 0.8123 | 0.9988 | 0.8677 | 0.7664 | 0.9998 | 0.9512 |

When comparing QUARTZ-GEN to QUARTZ-DL, disc segmentation performance slightly improved while cup segmentation performance slightly decreased. For UKBB, disc F1 increased from 0.9569 to 0.9599 and cup F1 decreased from 0.8469 to 0.8404. This disc improvement and cup decline pattern was consistent for the CLSA and NEL DESP datasets. Figure 5 shows a visual comparison of F1 performance.

The detection rate for optic disc localisation is summarised in Table 6, evaluated on an additional 300 images for the UKBB, CLSA, and NEL DESP datasets to increase the number of test images. Across the six internal datasets, QUARTZ-GEN achieved consistently high performance and low variability, with detection rates ranging from 0.9933 to 1.0000. On the external EPIC-Norfolk dataset, it attained a perfect detection rate of 1.0000. In comparison with QUARTZ-DL, QUARTZ-GEN showed slight improvements. Table 6 also reports the mean absolute error (MAE) for vCDR.

**Table 6:** Optic disc localisation and the MAE for vCDR. (i) denotes internal dataset; (e) denotes external dataset.

| Method | Dataset | Detection Rate | MAE for vCDR |
| --- | --- | --- | --- |
| QUARTZ-DL | UKBB (i) | 0.9933 | 0.0560 |
| QUARTZ-DL | CLSA (i) | 0.9900 | 0.0378 |
| QUARTZ-DL | NEL DESP (i) | 0.9900 | 0.0651 |
| QUARTZ-GEN | UKBB (i) | 0.9967 | 0.0591 |
| QUARTZ-GEN | CLSA (i) | 0.9933 | 0.0389 |
| QUARTZ-GEN | NEL DESP (i) | 0.9933 | 0.0700 |
| QUARTZ-GEN | REFUGE (i) | 1.0000 | 0.0382 |
| QUARTZ-GEN | PAPILA (i) | 1.0000 | 0.0656 |
| QUARTZ-GEN | Chákşu (i) | 1.0000 | 0.0367 |
| QUARTZ-GEN | ORIGA (e) | 1.0000 | 0.0696 |
| QUARTZ-GEN | EPIC-Norfolk (e) | 1.0000 | 0.0538 |

Table 7 summarises the image-quality classification performance. QUARTZ-GEN shows consistently strong and stable results across the three internal datasets. The F1 scores ranged from 0.9412 to 0.9655, with a mean F1 of 0.9525 ± 0.0100 and a CV of 1.05%. Its performance is lower on the external EPIC-Norfolk dataset, though it still attains a respectable F1 score of 0.9197. See Figure 4 for a visual comparison of F1 performance. When comparing QUARTZ-DL to QUARTZ-GEN, the UKBB F1 score decreases from 0.9750 to 0.9412, with declines observed for the CLSA and NEL DESP datasets (See Figure 5). Nevertheless, overall performance remains relatively high.

**Table 7:** Performance of image quality classification, positive class = inadequate. (i) denotes internal dataset; (e) denotes external dataset.

| Method | Dataset | Sensitivity | Specificity | Precision | NPV | Accuracy | F<sub>1</sub> Score | AUC ROC | AUC PR |
| --- | --- | --- | --- | --- | --- | --- | --- | --- | --- |
| QUARTZ-DL | UKBB (i) | 0.9512 | 1.0000 | 1.0000 | 0.9876 | 0.9900 | 0.9750 | 0.9974 | 0.9922 |
| QUARTZ-DL | CLSA (i) | 0.9118 | 1.0000 | 1.0000 | 0.9822 | 0.9850 | 0.9538 | 0.9945 | 0.9793 |
| QUARTZ-DL | NEL DESP (i) | 0.9737 | 0.9919 | 0.9867 | 0.9840 | 0.9850 | 0.9801 | 0.9950 | 0.9937 |
| QUARTZ-GEN | UKBB (i) | 0.9524 | 0.9810 | 0.9302 | 0.9873 | 0.9750 | 0.9412 | 0.9974 | 0.9902 |
| QUARTZ-GEN | CLSA (i) | 0.9063 | 1.0000 | 1.0000 | 0.9825 | 0.9850 | 0.9508 | 0.9931 | 0.9715 |
| QUARTZ-GEN | NEL DESP (i) | 0.9722 | 0.9766 | 0.9589 | 0.9843 | 0.9750 | 0.9655 | 0.9946 | 0.9924 |
| QUARTZ-GEN | FQS (e) | 0.9084 | 0.9711 | 0.8767 | 0.9791 | 0.9595 | 0.8923 | 0.9843 | 0.9554 |
| QUARTZ-GEN | EPIC-Norfolk (e) | 0.9248 | 0.9677 | 0.9147 | 0.9717 | 0.9560 | 0.9197 | 0.9919 | 0.9809 |

Results of the camera-level repeatability assessment are shown in Table 8. Across the 32 cameras, the within-participant vessel width measurements ranged from 87.26 to 97.79 microns and had a CV of 3.49%, indicating low camera-related variability when ‘participant’ is held constant. In contrast, the between-participant measurements showed a much wider range of 70.08 - 102.42 microns and a moderate CV of 6.59%, reflecting larger variation caused by effects of both biological differences and camera differences.

**Table 8:** Within-participant and between-participant vessel width variability. Measures are based on image-level mean vessel widths (in microns).

| Sample Group | No. | Vessel Type | Mean ($\mu\text{m}$) | SD ($\mu\text{m}$) | CV (%) | Range ($\mu\text{m}$) |
| --- | --- | --- | --- | --- | --- | --- |
| 1 participant | 32 images,<br>32 cameras | All vessels | 93.0983 | 3.2482 | 3.4890 | 87.2630 - 97.7871 |
|  |  | Arterioles | 86.6614 | 2.6138 | 3.0161 | 81.5232 - 91.3869 |
|  |  | Venules | 99.4687 | 4.7991 | 4.8248 | 88.6348 - 108.1782 |
| 100 participants | 100 images,<br>multiple cameras | All vessels | 84.4722 | 5.5661 | 6.5893 | 70.0799 - 102.4236 |
|  |  | Arterioles | 79.9090 | 6.6760 | 8.3546 | 60.5801 - 99.0394 |
|  |  | Venules | 89.0295 | 6.4742 | 7.2720 | 76.5864 - 116.1512 |

The results from processing the entire datasets are detailed in Table 9, reporting the numbers of images that have useful information extracted for use in epidemiological studies. Participant consent withdrawals result in a small reduction in the number of available UKBB retinal images. The National Health Service (NHS) NEL DESP dataset has expanded to 4,356,007 images (now exclusively retinal images, with non-retinal images excluded). Processing times have improved due to threading of the CPU tasks, now taking 1.005 seconds on average to process a single UKBB image using an Intel i9-12900KF × 24 threads and an RTX 4090 with 24 GB VRAM and 32 GB system RAM (for context, training each deep learning model required up to 6 hours on the same hardware). The QUARTZ online tool operates on more limited computational resources. Although a direct comparison with other systems is beyond the scope of this work, the model demonstrates performance that is comparable to current state-of-the-art systems. Reported segmentation results from current leading approaches [8,9] are provided for context in Table 10.

**Table 9:** Processing entire datasets.

| Method | Dataset | No. of Images | Adequate Image Quality |
| --- | --- | --- | --- |
| QUARTZ-DL | UKBB | 175,764 | 132,293 (75.27%) |
| QUARTZ-DL | CLSA | 106,506 | 88,155 (82.77%) |
| QUARTZ-DL | NEL DESP | 1,175,423 | 803,555 (68.36%) |
| QUARTZ-GEN | UKBB | 175,620 | 128,734 (73.30%) |
| QUARTZ-GEN | NEL DESP | 4,356,007 | 3,908,462 (89.73%) |
| QUARTZ-GEN | EPIC | 16,044 | 11,780 (73.42%) |

**Table 10:**
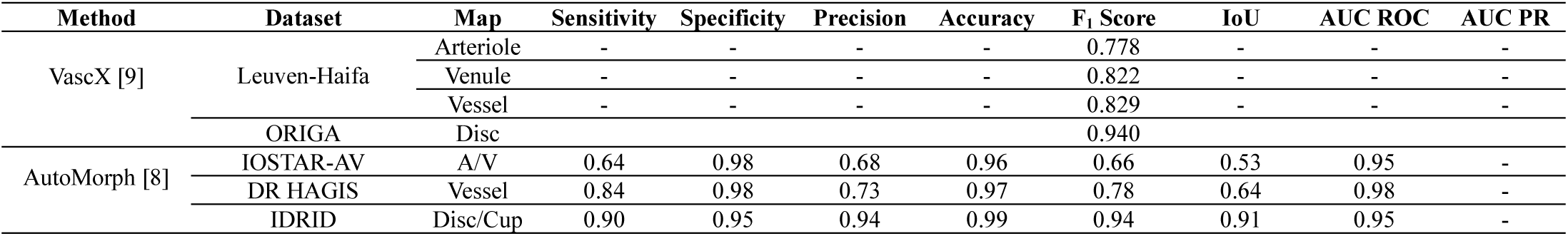
Reported performances of alternative vasculometry approaches: A/V segmentation (prior to post-processing) and optic disc/cup segmentation.

## (5) Discussion and Conclusion

QUARTZ is a fully automated retinal vasculometry system that extracts quantitative vascular measures at scale for use in epidemiological research and disease risk prediction. In this paper, a generalisation-focused version of QUARTZ has been demonstrated to be robust, producing predominantly high performance and low variability across multiple diverse datasets. These datasets include variations in population demographics, disease status, and differences in imaging environments such as camera systems, FOV angles, resolution, image quality, and pupil dilation status. Widefield images were avoided to focus the system to a FOV range of 30 - 60°.

Low variability was exhibited by the deep learning modules, with predominantly low CV across multiple internal datasets, highlighting the system’s stability. Values were 2.97%, 2.40%, and 2.12% for arteriole, venule, and vessel segmentation, respectively; 1.13% and 4.52% for optic disc and optic cup segmentation; and 1.05% for image quality classification. Variability was also low for the resulting A/V segment-level decisions and optic disc localisation. The 4.52% value for optic cup segmentation was the only notable deviation, yet it still indicates low-to-moderate variability. This deviation is due to the performance on the PAPILA dataset. Although a high quality and carefully curated dataset, optic cup annotation is inherently subjective, and parts of the cup from which vessels emerge were not annotated as cup, likely explaining the lower performance value. Despite this, a good mean F1 score of 0.8520 for optic cup segmentation was achieved across all datasets. Importantly, the modules maintained robust performance even in challenging scenarios, such as images with dense choroidal vasculature, irregular optic cup shapes, laser scars, or various retinal pathologies, demonstrating consistent reliability across diverse cases.

QUARTZ also generalised effectively to the external EPIC-Norfolk dataset, maintaining high performances within a range similar to that observed on the internal datasets. Results on the additional external datasets (Fundus-AVSeg, ORIGA, and FQS) showed broadly similar performance, indicating progress toward the system’s goal of enhanced external generalisation.. These findings suggest that the system’s generalisation- focused design has translated into performance stability across heterogeneous external cohorts. However, an even broader evaluation would be needed to support fully a claim to describe the system as domain generalisable Whilst comparison to other systems is not our intention here, QUARTZ performance compares well with current state-of-the-art systems.

Traditionally, moving a system toward a more generalised approach can improve cross-dataset robustness, though this may come with a reduction in performance compared with a dataset-tailored approach. In the case of QUARTZ, however, this is not the direct comparison being made. Rather, the dataset-tailored version was constrained by having access to relatively limited training data, while the generalisation-focused version benefits from a much larger pool of data, enabling longer training and the learning of richer feature representations. This advantage was reflected in improved performance for A/V and optic disc segmentation. In contrast, no similar gains were observed for optic cup segmentation, likely due to the variability in the optic cup annotation amongst the datasets, or for image quality classification, where the issues can be highly dataset specific.

This latest version of QUARTZ improves A/V segmentation performance, building on the substantial enhancements of previous versions, and continues to increase the number of small vessels represented in the segmented vessel distribution. While this provides a richer source of information, it may also affect previously reported associations that focused primarily on larger vessels. In fact, associations or predictions derived from image-level averages that predominantly reflect larger (wider) vessel segments may differ when image-level averages are instead calculated using more comprehensive vascular tree measures that include a higher proportion of smaller (narrower) vessel segments. For analyses specifically targeting large retinal vessels rather than pre- and post-capillaries, it may therefore be beneficial to exclude vessel segments with smaller widths.

While current datasets enabled a limited study of repeatability, QUARTZ demonstrates relatively low variation in the camera-level repeatability assessment with a within-participant CV of 3.49%. However, this is approximately half of the between-participant CV (6.59%), indicating that camera related differences are a meaningful component of the total variability. This suggests that while QUARTZ measurements are broadly consistent across devices, camera heterogeneity does introduce measurable noise. Nevertheless, because this noise is appreciably smaller than the overall between-person variability, the underlying biological signal remains detectable in large epidemiological studies. In this context, QUARTZ is sufficiently robust for identifying population-level associations. It should be noted that this assessment used image-level mean vessel width, which although widely employed in epidemiological research, can be sensitive to variations in the captured vessel distribution. Such variability may arise from differences in image resolution or the camera’s ability to capture details, as well as the operator’s skill in achieving sharp focus and correctly centring on the macula.

A limitation of this study is that QUARTZ’s image quality score is based on A/V segmentation rather than optic disc/cup segmentation, enabling effective use of vessels even in partially illuminated images. However, optic disc and cup measurements can be affected by poorly illuminated optic discs. For epidemiological studies using vCDR or optic disc/cup size outputs, a separate quality assessment focused on optic disc clarity is recommended. Future work will introduce metrics specifically for optic disc/cup segmentation quality.

Alternatively, optic disc/cup measurements that deviate substantially from the population mean can be excluded.

While QUARTZ is intended for retinal images with a FOV range of 30–60°, evaluation was limited by available annotated datasets. A/V classification was evaluated across the full range, whereas optic disc/cup segmentation and image quality classification were only evaluated on 30–45° images. Images outside these ranges were inspected qualitatively, but full metric-based evaluation was not performed; therefore, performance on such images remains unverified and should be interpreted accordingly.

An additional limitation is that although the balanced sampling strategy mitigates dataset dominance during training, some smaller datasets may still contribute to class imbalance, leading to potential underrepresentation of certain demographic groups and pathologies, which could impact generalisation.

In conclusion, this work aimed to develop a generalisation-focused version of QUARTZ capable of robust operation across a broad range of retinal datasets, enabling wider adoption and reproducible biomarker research. Training QUARTZ on multiple diverse datasets has produced a system that performs robustly across varied data. Its performance is comparable to dataset-specific versions and exceeds them in certain modules.

Epidemiologists in our team have confirmed that established associations remain robust with the latest version of QUARTZ; detailed analyses will be reported separately. To further support reproducible biomarker research, an online tool has been released to broaden access. QUARTZ has also been applied to large-scale retinal datasets and has analysed over 4 million images from within the NHS. These retinal vasculometry outputs will serve as a valuable resource for future epidemiological or other clinical studies. In addition, the new public CHASE-AV database is introduced, providing an additional resource for the research community.

## Collaborators

The UK Biobank Eye and Vision Consortium: the list of members is available from the consortium website (http://www.ukbiobankeyeconsortium.org.uk/people).

The EPIC-Norfolk Investigators: the list of members is available from the study website (https://www.epic-norfolk.org.uk/about-epic-norfolk/people).

The Artificial Intelligence & Automated Retinal Image Analysis Systems (ARIAS) Research Group: John Anderson, Sarah Barman, Louis Bolter, Ryan Chambers, Lakshmi Chandrasekaran, Umar Chaudhry, Emily Y Chew, Catherine Egan, Jiri Fajtl, Frederick L Ferris, Aroon D Hingorani, Aaron Y Lee, Abraham Olvera-Barrios, Christopher G Owen, Paolo Remagnino, Alicja R Rudnicka, Royce Shakespeare, Reecha Sofat, Adnan Tufail, Alasdair N Warwick, Charlotte Wahlich, Roshan Welikala, Kathryn Willis, Yue Wu.

## Acknowledgements

This research has been conducted using the UK Biobank resource under application number 522. The UK Biobank Eye and Vision Consortium is supported by funding from The Special Trustees of Moorfields Eye Hospital NHS Foundation Trust, and at the NIHR Biomedical Research Centre at Moorfields Eye Hospital and UCL Institute of Ophthalmology.

This research was made possible using the data/biospecimens collected by the Canadian Longitudinal Study on Aging (CLSA). Funding for the Canadian Longitudinal Study on Aging (CLSA) is provided by the Government of Canada through the Canadian Institutes of Health Research (CIHR) under grant reference: LSA 94473 and the Canada Foundation for Innovation, as well as the following provinces, Newfoundland, Nova Scotia, Quebec, Ontario, Manitoba, Alberta, and British Columbia. This research has been conducted using the CLSA Baseline Comprehensive Dataset version 7.0 and Follow-up 1 Comprehensive Dataset version 4.0 and Baseline Retinal Scans under Application Number 2209017. The CLSA is led by Drs. Parminder Raina, Christina Wolfson and Susan Kirkland. The time and commitment of the participants to the CLSA study platform is gratefully acknowledged, without whom this research would not be possible. The opinions expressed in this manuscript are the author’s own and do not reflect the views of the Canadian Longitudinal Study on Aging. Data are available from the Canadian Longitudinal Study on Aging (www.clsa-elcv.ca) for researchers who meet the criteria for access to de-identified CLSA data.

The EPIC-Norfolk study (DOI 10.22025/2019.10.105.00004) has received funding from the Medical Research Council (MR/N003284/1 and MC-UU_12015/1) and Cancer Research UK (C864/A14136). The genetics work in the EPIC-Norfolk study was funded by the Medical Research Council (MC_PC_13048). We are grateful to all the participants who have been part of the project and to the many members of the study teams at the University of Cambridge who have enabled this research.

Collection of data from the North East London Diabetic Eye Screening Programme (NEL DESP), part of Homerton Healthcare NHS Foundation Trust, and image analysis work was funded by the Wellcome Trust Collaborative Award (224390/Z/21/Z), and conducted by the ARIAS Research Group.

## Data availability

The DESP dataset is not publicly available due to patient privacy constraints and NHS data governance requirements. UK Biobank data are available to approved researchers via the UK Biobank Access Management System (https://www.ukbiobank.ac.uk/enable-your-research/apply-for-access). CLSA data are available from www.clsa-elcv.ca for researchers who meet the criteria for access to de-identified CLSA data. EPIC-Norfolk data are available from https://www.epic-norfolk.org.uk for researchers who meet the criteria for access.

FOREVER data is available https://forever.ku.dk/ for researchers who meet criteria for access. All public benchmark datasets (Leuven-Haifa, MESSIDOR, HRF-AV, LES-AV, Chákṣu, CHASE-DB1, CHASE-AV, FQS, Fundus-AVSeg, REFUGE, PAPILA, and ORIGA, RITE) are available at their respective repositories cited in the text.

## Ethics approval and patient consent

The DESP dataset comprises clinical images collected as part of routine NHS diabetic eye screening. All data were managed according to UK NHS information governance requirements, in accordance with the principles of the Data Protection Act 2018. Methodologies adhere to the Homerton Healthcare NHS Foundation Trust Security Policy and Information Governance Policy, and General Data Protection Regulation principles relating to the processing of personal data. The study has been approved by the Health Research Authority (IRAS project ID 265637). The NHS Health Research Authority toolkit identified that research ethics approval was not required for this project, as all data are pseudonymised and presented in aggregate form. A Data Protection Impact Assessment was submitted to the Homerton Healthcare NHS Foundation Trust Information Governance Lead and approved by the information governance team in December 2022. The UK Biobank study was carried out following the principles of the Declaration of Helsinki and the Research Governance Framework for Health and Social Care. The UK Biobank study was approved by the North West Multi-Centre Research EthicsCommittee (11/NW/03820). All participants gave written, informed consent.

## Permission to reproduce material from other sources

Images shown within the manuscript from datasets used have been acknowledged within the text and captions (Leuven-Haifa, MESSIDOR, HRF-AV, LES-AV, Chákṣu,, REFUGE, PAPILA, and ORIGA)

